# A Multimodal Evaluation of Transcranial Photobiomodulation in Mild Cognitive Impairment: Cognitive, Metabolic, and Neuroimaging Outcomes of a Pilot Randomized Controlled Trial

**DOI:** 10.1101/2025.08.19.25333989

**Authors:** Neda Rashidi-Ranjbar, Nathan W. Churchill, Mirjana Jerkic, Reza Zomorrodi, Ori Rotstein, Raphael Schneider, Ana C. Andreazza, Tarek K. Rajji, Simon J. Graham, David G. Munoz, Luis Fornazzari, Lew Lim, Mireille Norris, Tom A. Schweizer, Corinne E. Fischer

**Affiliations:** Keenan Research Centre for Biomedical Science, Li Ka Shing Knowledge Institute, St. Michael’s Hospital, Toronto, ON, Canada; Temerty Centre for Therapeutic Brain Intervention, Centre for Addiction and Mental Health, Toronto, Ontario, Canada; Temerty Faculty of Medicine, Department of Surgery, University of Toronto, Toronto, ON, Canada; Temerty Faculty of Medicine, Division of Neurology, University of Toronto, Toronto, ON, Canada; Temerty Faculty of Medicine, Department of Psychiatry, University of Toronto, Toronto, ON, Canada; Department of Pharmacology & Toxicology, Mitochondrial Innovation Initiative, University of Toronto, Toronto, ON, Canada; Department of Psychiatry, UT Southwestern Medical Center, Dallas, Texas; Department of Medical Biophysics, University of Toronto, Toronto, Ontario, Canada; Physical Sciences Platform, Sunnybrook Research Institute, Sunnybrook Health Sciences Centre, Toronto, ON, Canada; Temerty Faculty of Medicine, Department of Laboratory Medicine and Pathobiology, University of Toronto, Toronto, ON, Canada; Vielight Inc, Toronto, ON, Canada; Temerty Faculty of Medicine, Department of Medicine, University of Toronto, Toronto, ON, Canada; Temerty Faculty of Medicine, Department of Neurosurgery, University of Toronto, Toronto, ON, Canada

## Abstract

**INTRODUCTION:** Mild cognitive impairment (MCI), a prodromal stage of Alzheimer’s disease and related dementias (ADRD), represents a critical window for intervention. Although mitochondrial dysfunction is increasingly implicated in neurodegeneration, most therapies target downstream protein aggregation. Transcranial photobiomodulation (tPBM) delivers near-infrared light to enhance mitochondrial respiration. We hypothesized that tPBM in MCI would be safe, feasible, and associated with improvements in cognition, mitochondrial function, and default-mode-network (DMN) functional connectivity (FC).

**METHODS:** We conducted a single-blind, randomized, sham-controlled pilot trial (NCT05563298) in adults ≥50 years with MCI. Twenty participants were randomized 1:1 to active or sham devices. Active devices delivered pulsed 810-nm light for 20 minutes per session; shams emitted light for 2 seconds. Stimulation targeted DMN hubs and the olfactory bulb. Participants self-administered treatment at home six days per week for six weeks.

**RESULTS:** Adherence was high (active 96.9%; sham 94.2%). Adverse events (Aes) were reported by 10 of 20 participants (4 active, 6 sham) No serious AE occurred. Compared with sham, active tPBM produced greater improvement in global cognition (MMSE; p=0.03) and episodic memory (CVLT-II long-delay recognition; p=0.02). Serum pyruvate and lactate increased with a reduced lactate-to-pyruvate ratio (p=0.007). DMN FC increased (p=0.014), and plasma IL-6 declined (p=0.02).

**DISCUSSION:** Home-based tPBM was safe, tolerable, and feasible, with strong adherence and mild AEs. Cognitive, metabolic, and network-level findings suggest enhanced mitochondrial efficiency and anti-inflammatory effects. These results support larger, double-blind multicenter trials to evaluate tPBM as a mitochondria-targeted therapy in early ADRD.

## 1. Background

Alzheimer’s disease and related dementias (ADRD) affect over 55 million people worldwide, with ∼10 million new cases annually^1^. By symptom onset, extensive synaptic and neuronal loss has already occurred, limiting treatment efficacy^2^. Anti-amyloid agents such as lecanemab provide modest benefit in early ADRD^3^ but are restricted by cost, infrastructure requirements, and risks of amyloid-related abnormalities^3^. As up to 45% of dementia cases may be preventable with early intervention^4^, scalable, noninvasive strategies for high-risk groups such as mild cognitive impairment (MCI) are urgently needed.

MCI is defined by measurable decline, typically in episodic memory, without major functional loss^5^. About 15% progress to dementia within two years, and one-third to ADRD within five^5^. Mitochondrial dysfunction is recognized as an early hallmark of ADRD^6^. In fact, reduced mitochondrial cytochrome c oxidase activity and elevated reactive oxygen species drive redox imbalance and accelerate amyloid and tau pathology^6^. Yet most therapies continue to target amyloid, tau, or inflammation^7^. The scarcity of mitochondria-directed approaches underscores the need for novel interventions that directly address this critical mechanism in ADRD pathogenesis.

Transcranial photobiomodulation (tPBM) is a non-invasive approach that delivers near-infrared (NIR) light (700–1100 nm) to stimulate cytochrome c oxidase (CCO) and enhance mitochondrial respiration^8^. Near-infrared spectroscopy studies confirm that these wavelengths coincide with CCO’s absorption peak, enabling efficient photon capture and modulation of oxidative metabolism^8^. tPBM increases ATP production, reduces reactive oxygen species, and promotes nitric oxide release^9^. In Alzheimer’s disease (AD) mouse models, such as 5×FAD (five familial Alzheimer’s disease mutations) and amyloid precursor protein/presenilin-1 (APP/PS1), using wavelengths near 808 ± 10 nm delivered either continuously or pulsed at 100 Hz, and in one study, using 670 nm light applied in a 90-second cycle, reduced Aβ burden, preserved synaptic integrity, and improved memory^10,11^. Early human trials in MCI^12^ and AD^12–14^ reported gains in attention, executive function, memory, cerebral blood flow^12^, and default mode network (DMN) functional connectivity (FC)^14^, using continuous 610 ± 10 nm light^12^ or pulsed 810 nm light at 10 Hz^13^ and 40 Hz (50% duty cycle)^14^. However, these studies were few and generally limited by small samples and lack of sham controlled^12–14^.

To address these gaps, we conducted a single-blind, randomized, sham-controlled feasibility trial of tPBM in individuals with MCI (n = 20; NCT05563298). The primary endpoint was change in cognition—including global function, episodic memory, and processing speed—from baseline to week 7. Secondary endpoints examined mechanistic effects on serum lactate, pyruvate, and the lactate/pyruvate ratio^15^, neurometabolite ratios in the posterior cingulate cortex (PCC)—a DMN hub and early site of metabolic decline in ADRD^16^—DMN FC, and cerebral blood flow. Exploratory outcomes included blood biomarkers of neurodegeneration and inflammation, MRI markers of atrophy, neuropsychiatric symptoms, and sleep quality. By integrating cognitive, biomarker, and imaging measures, this pilot trial evaluated feasibility, safety, preliminary efficacy, and mechanistic pathways to guide future large-scale RCTs of tPBM in early cognitive decline.

## 2. Methods

### 2.1. Participants and Study Design

Between March 2023 and December 2024, we enrolled twenty community-dwelling adults aged 60–85 years with mild cognitive impairment (MCI) due to Alzheimer’s disease (NCT05563298). Diagnoses were established by consensus between a geriatric psychiatrist and a behavioral neurologist according to Petersen and Albert et al. (2011)^17^ criteria for MCI due to AD, incorporating clinical judgment and supportive structural MRI and single-photon emission computed tomography (SPECT) findings, and consistent with NINCDS-ADRDA guidelines for underlying Alzheimer’s pathology^18^. Participants were referred from the St. Michael’s Hospital Memory Clinic or recruited through community advertisements. All procedures were conducted at St. Michael’s Hospital under Research Ethics Board approval, with written informed consent obtained from all participants. Inclusion criteria required a subjective cognitive complaint, MoCA score of 19–25 with memory impairment, Clinical Dementia Rating (CDR) of 0.5 with preserved daily function, at least eighth-grade education, and English fluency. Exclusion criteria included MRI or venipuncture contraindications, major psychiatric or neurological illness, uncontrolled systemic disease, history of stroke or seizure, photosensitivity, major anticoagulant use, or inability to provide informed consent.

### 2.2. Study Design and Sample Size

This single-blind randomized sham-controlled trial included 10 participants per arm, consistent with recommendations for pilot studies to inform design of a definitive trial^19^. Participants were randomized 1:1 (block size of four) to active or sham tPBM. Allocation was managed by a single unblinded coordinator who performed screening, follow-up, and assessments, while participants were blinded to condition, constituting a single-blind, sham-controlled design. The study flow is summarized in Figure 1 (CONSORT diagram).

**Figure 1.**
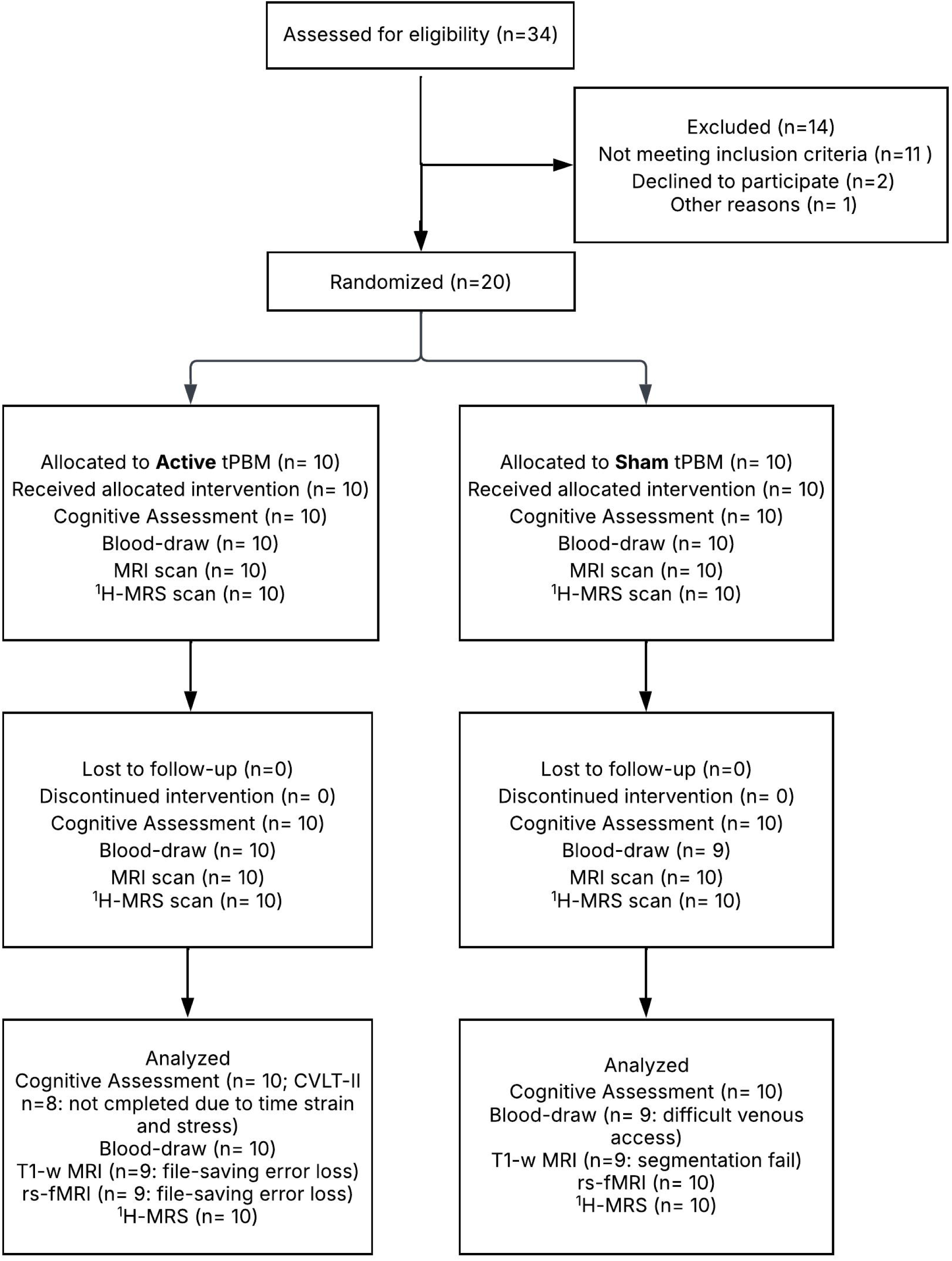
CONSORT participant flow. Diagram shows screening, randomization to active vs sham tPBM, retention, and modality-specific analysis sets. Reasons for missing data are annotated in-figure (data-saving error during acquisition; segmentation failure due to pituitary adenoma; ASL parameter mismatch; two CVLT-II noncompletions; one missed post-blood draw). Abbreviations: tPBM, transcranial photobiomodulation; rs-fMRI, resting-state fMRI; ¹HMRS, proton MR spectroscopy; pCASL, pseudo-continuous arterial spin labeling.

### 2.3. Intervention

Participants received a portable Class II research device (Health Canada ITA #347303) containing five 810 nm LEDs targeting the medial prefrontal, posterior cingulate, temporoparietal, and intranasal regions (Figure 2). Active devices emitted pulsed near-infrared light at 40 Hz (50% duty cycle), delivering 75–100 mW/cm² at the scalp and 25 mW/cm² intranasally, with heat generation <0.5 °C. Sham devices were visually identical but deactivated after a brief initial pulse to preserve blinding. All devices complied with Class II Health Canada safety standards.

**Figure 2.**
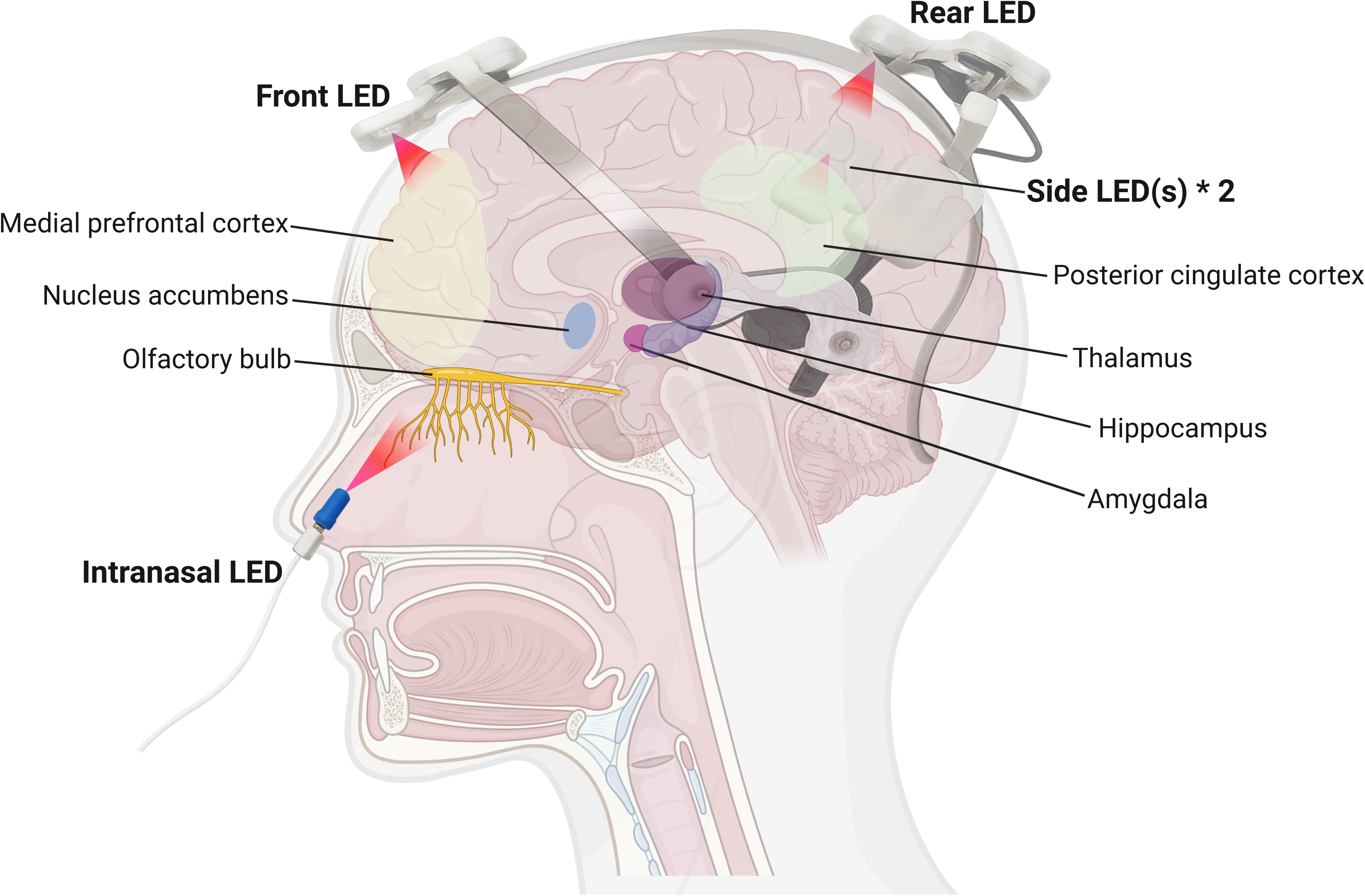
Schematic of the Transcranial photobiomodulation (tPBM) headset and projected light paths illustrates the five LED emitters used in the present study. Red cones depict the approximate spread of five LED emitters. The frontal LED, positioned at the front hairline, targets the medial prefrontal cortex; the posterior LED on the crown illuminates the posterior cingulate cortex; the two lateral LEDs (only one visible in this view) fall naturally into position without manual adjustment, covering the temporoparietal cortices; and an intranasal LED, inserted in the nostril, targets the olfactory bulb. Subcortical structures are highlighted for reference. Illustration created with BioRender.com.

**Table 1.** Demographic characteristics of the active and sham groups.

Based on Monte Carlo modeling and optical measurements, approximately 1–3% of incident 810 nm light reaches cortical tissue after passing through scalp and skull, corresponding to ∼0.75–3 mW/cm² at the cortical surface^20^. Participants were instructed to part their hair at stimulation sites to minimize attenuation and ensure consistent scalp contact. Nonetheless, hair thickness and color may variably attenuate transmission, representing an unavoidable source of inter-individual variability.

At baseline, the coordinator conducted a 30-minute in-person training session on device placement, cleaning, and diary use. Participants completed 20-minute sessions six days per week for six weeks (36 total), consistent with prior feasibility and safety studies^21^ and corresponding to an estimated energy density of 45–60 J/cm² at the scalp, within the established therapeutic range for cortical PBM. The intranasal emitter delivered approximately 15 J/cm² (25 mW/cm² × 50 % duty cycle × 1200 s).

Devices were typically used at bedtime to standardize timing, though minor variability in session schedule or pre-treatment state was permitted.

Adherence and safety were monitored through diaries, weekly calls, and an adverse-event log. Calls confirmed session completion, addressed device issues, and documented side effects.

Compliance was defined as ≥30 of 36 sessions (∼83%), allowing one missed session per week, consistent with the 80% adherence threshold validated in prior systematic reviews^22^.

### 2.4. Outcome Measures

To comprehensively evaluate treatment effects while minimizing participant burden, the study incorporated cognitive, biomarker, and neuroimaging outcomes that together capture multiple domains relevant to MCI. All post-treatment assessments were completed within three days (±2 days) of the final stimulation session. Cognitive testing and blood draws were performed on the same day, and MRI/¹H-MRS scans were obtained either the same day or within the following two days. A follow-up visit one month after treatment assessed the durability of effects.

#### 2.4.1. Cognitive Function Assessments (n = 10 active, n = 10 sham)

Cognitive testing (n=10 active, n=10 sham) was conducted at baseline and week 7 by the same trained coordinator. The battery assessed global cognition, episodic and recognition memory, processing speed, sleep quality, and neuropsychiatric symptoms. Global cognition was measured with the MMSE^23^. Processing speed was assessed using the TMT-A, TMT-B, and the B/A ratio^24^. Episodic memory was evaluated with the CVLT-II, using alternate forms at post-test to minimize practice effects (Cohen’s d = –0.01–0.18)^25^. Outcomes included total recall (Trials 1– 5), delayed free recall, and long-delay recognition hits. Sleep quality and neuropsychiatric symptoms were assessed using the Pittsburgh Sleep Quality Index (PSQI)^26^ and Mild Behavioral Impairment Checklist (MBI-C)^27^, respectively.

#### 2.4.2. Blood-Based Biomarkers (n = 10 active, n = 9 sham)

Blood samples were collected immediately after cognitive testing at baseline and week 7. Whole blood was drawn into two 10 mL vacutainer tubes—one containing K EDTA and one without anticoagulant, which was allowed to clot upright at room temperature for 30–45 minutes. EDTA and serum tubes were then centrifuged at 2000 x g for 25 minutes at room temperature or at 4°C, respectively (Sorvall ST16 Plus centrifuge, ThermoFisher Scientific, Markham, ON, Canada) using ClickSeal biocontainment lids.. Plasma and serum were aliquotted under sterile conditions in a biosafety cabinet, inspected for turbidity, and re-centrifuged if needed. All aliquots were stored in cryovials at –80°C within two hours of collection.

##### 2.4.2.1. Serum Metabolomics Analysis

NMR spectroscopy were performed by the Metabolomics Innovation Centre (TMIC, University of Alberta; https://metabolomicscentre.ca/service/nmr-analysis/), with a primary focus on lactate and pyruvate as markers of mitochondrial function. To remove high-molecular-weight proteins and lipoproteins, samples were deproteinized using 3 kDa cut-off centrifugal filters (Amicon Microcon YM-3), pre-rinsed to eliminate residual glycerol^28^. Samples were centrifuged at 10,000 rpm for 20 minutes, and filtrates were inspected for membrane integrity. If volumes were below 250 µL, 150 mM KH□PO□ buffer (pH 7.0) was added to reach 173.5 µL. A standard buffer (54% D□O, 5.84 mM DSS, 5.84 mM 2-chloropyrimidine-5-carboxylate, 0.1% NaN□) was then added (46.5 µL). ¹H-NMR spectra were acquired at 25°C using a Bruker Avance III 700 MHz spectrometer with a 5 mm HCN cryoprobe, applying the NOESY pre-saturation sequence (noesy1dpr) for quantitative accuracy^29^. The DSS methyl signal was used as an internal standard (0 ppm). Spectra were processed and quantified using an in-house version of MAGMET software^29,30^, which fits known metabolite signatures from a custom database.

##### 2.4.2.2. Plasma Biomarkers Analysis

Exploratory plasma markers of inflammation (IL-1β, IL-10, IL-6, TNF-α)^31^, synaptic plasticity (BDNF)^31^, and neurodegeneration (p-Tau217, NfL)^31^ were quantified using the Ella microfluidic platform (ProteinSimple, Bio-Techne) and the Simoa® ALZpath p-Tau217 assay (Quanterix). Plasma aliquots were thawed once and centrifuged at 10,000 × g for 10 minutes at 4°C, to remove any particles that might be present in the samples. Biomarker and cytokine concentrations (pg/mL) were measured using the Ella platform (ProteinSimple, Bio-Techne, New York, USA), an automated microfluidic immunoassay system, with V5-generation single and multiplex cartridges. Assays followed the manufacturer’s technical guidelines^32^. Phosphorylated tau-217 was measured separately using the Simoa® ALZpath p-Tau 217 assay (Quanterix).

#### 2.4.3. Neuroimaging Acquisition

MRI was acquired at baseline and week 7 on a 3 T Siemens Skyra with a 64-channel head coil, including 3D T1-weighted (MPRAGE), FLAIR, resting-state fMRI, single-voxel ¹H-MRS (posterior cingulate cortex), and pseudo-continuous ASL (pCASL) for cerebral blood flow.

One participant’s T1 and rs-fMRI were lost to a file-saving error (¹H-MRS was retained); another participant’s T1 failed segmentation because of a pituitary adenoma and was excluded from structural analyses; and one ASL dataset was excluded for acquisition parameter mismatch. **F**inal analyzable samples were: structural MRI 9 active/9 sham, rs-fMRI 9 active/10 sham, PCASL 9 active/9 sham, and ¹H-MRS 10 active/10 sham

##### 2.4.3.1. rs-fMRI (n = 9 active, n = 10 sham)

Resting-state functional MRI was acquired using 2D multi-slice T2*-weighted echo planar imaging (EPI; TE/TR = 30/2000 ms, flip angle = 70°, 36 oblique-axial slices, 200 × 200 mm field of view, 64 × 64 matrix, 3.5 mm slice thickness with 0.5 mm gap, 3.1 × 3.1 mm in-plane resolution, 2442 Hz/px bandwidth), producing 270 brain volumes (9 minutes total). Participants were instructed to lie still with eyes closed and not focus on anything in particular, and they confirmed verbally that they were awake and not drowsy before and after scanning. Data were processed using a hybrid pipeline incorporating ANTs (Advanced Normalization Tools; http://stnava.github.io/ANTs), AFNI (Analysis of Functional NeuroImages; https://afni.nimh.nih.gov), FSL (FMRIB Software Library; https://fsl.fmrib.ox.ac.uk/fsl), and inhouse scripts^36^. Functional images were non-linearly normalized to MNI152 space and resampled to 3 mm isotropic resolution. Spatial smoothing was applied using a 6 mm full-width at half-maximum Gaussian kernel. Subsequent preprocessing included linear detrending, regression of six motion parameters, and removal of nuisance signals from white matter (WM) and cerebrospinal fluid (CSF) time series. To ensure data quality, frames with framewise displacement >0.5 mm were identified and censored. Participants with >20% of volumes flagged for motion would have been excluded, but no participants met this threshold, and mean framewise displacement did not differ significantly between groups.

We applied the Schaefer 200-parcel cortical atlas (Yeo’s seven networks) ^37^ plus the Tian 16region subcortical atlas (eighth network)^38^ to extract mean time series from 216 ROIs. Fisher-ztransformed Pearson correlations were computed between all region of interest (ROI) pairs, producing 216 × 216 connectivity matrices. Within-network connectivity was assessed as the mean z-value of all ROI pairs in a given network; between-network connectivity was assessed as the mean z-value between ROIs of two distinct networks.

rs-fMRI data were preprocessed using an in-house pipeline^36^ using AFNI, FSL, and ANTs with slice-timing and motion correction, MNI152 normalization, nuisance regression, band-pass filtering (0.01–0.1 Hz), and 6 mm FWHM smoothing. Frames with >0.5 mm framewise displacement were censored; none exceeded 20%. Time series were parcellated using the Schaefer 200-parcel cortical atlas^37^ and Tian 16-region subcortical atlas^38^, combined into a 216region mask. Mean time series were extracted using *fslmeants, and* Fisher z-transformed Pearson correlations were computed to quantify within- and between-network connectivity, including the default mode network (DMN).

##### 2.4.3.2. ¹H-MRS (n = 10 active, n = 10 sham)

Single-voxel proton magnetic resonance spectroscopy (¹H-MRS) was performed in the posterior cingulate cortex (voxel size = 20 × 20 × 20 mm³) using a Point RESolved Spectroscopy (PRESS) sequence (TE/TR = 30/2000 ms, flip angle = 90°, 80 water-suppressed and 4 unsuppressed averages, 1024 points, 1200 Hz bandwidth). Voxels were manually placed on the mid-sagittal plane of the posterior cingulate cortex, centered above the corpus callosum. To ensure reproducibility across sessions, a screenshot of baseline voxel placement was saved for each participant and used to guide placement at the post-treatment scan. Visual inspection confirmed identical voxel overlap between baseline and post-treatment acquisitions.

^1^H-MRS spectra were fitted with TARQUIN v4.3.10 (https://tarquin.sourceforge.net). Spectra with Cramér–Rao Lower Bounds < 20% were retained; none were excluded. Metabolite concentrations (N-acetylaspartate [NAA], choline [Cho], myo-inositol [mI], and lactate [Lac]) were expressed as ratios to creatine (e.g., NAA/Cr, Lac/Cr).

##### 2.4.3.3. T1-weighted imaging (n = 9 active, n = 9 sham)

High-resolution structural imaging included a 3D T1-weighted Magnetization Prepared Rapid Acquisition Gradient Echo (MPRAGE) sequence (TI/TE/TR = 1100/4.37/2500 ms, flip angle = 7°, 192 sagittal slices, 256 × 256 matrix, 1.0 mm slice thickness, 1.0 × 1.0 mm in-plane resolution, 140 Hz/px bandwidth), and a 3D Fluid Attenuated Inversion Recovery (FLAIR) sequence (TI/TE/TR = 1600/388/5000 ms, 192 sagittal slices, 256 × 256 matrix, 1.0 mm slice thickness, 1.0 × 1.0 mm in-plane resolution, 751 Hz/px bandwidth).

T1-weighted were processed with FreeSurfer (Longitudinal) v6.0 with visual quality control (QC) and manual correction. QC procedures included detailed visual inspection of cortical and subcortical segmentations to ensure accuracy. For each participant, change scores (Δ = week 7 – baseline) were calculated for bilateral volumes of six subcortical bilateral regions (hippocampus, amygdala, thalamus, caudate, putamen; n = 12) using the “aseg” parcellation^33^, and for bilateral cortical thickness across eighteen bilateral (n = 36) Desikan–Killiany regions^34,35^

##### 2.4.3.4. Arterial Spin Labeling (n = 9 active, n = 9 sham)

Pseudo-continuous arterial spin labeling (PCASL) was acquired with TR/TE = 4930/36.8 ms, flip angle = 120°, 8 tag-control pairs, 1500 ms labeling duration, and 1800 ms post-labeling delay. Scans were acquired with 2.5 mm isotropic resolution over a 240 × 240 mm field of view, using 48 slices and background suppression. A 3D turbo gradient spin-echo (TGSE) readout was used with 2-fold in-plane acceleration via GRAPPA (2480 Hz/px bandwidth). A calibration scan was also acquired using identical parameters but without tagging pulses or background suppression, and with a TR of 7000 ms.

Pseudo-continuous arterial spin labeling (PCASL) data were processed using a custom pipeline that integrates in-house software with standard open-source packages, including including AFNI (https://afni.nimh.nih.gov), FSL (https://fsl.fmrib.ox.ac.uk/fsl), and ANTs (http://stnava.github.io/ANTs)^39^. Outlier volumes were identified and removed, and the remaining volumes were aligned to the corresponding M calibration scan. Cerebral blood flow (CBF) maps were estimated in mL/100g/min using a kinetic model adapted from ASLtbx^40^ with default parameters. The resulting CBF maps were normalized to MNI space, resampled to 3 mm isotropic resolution, and spatially smoothed using a 6 mm full-width at half-maximum (FWHM) Gaussian kernel. Next, we extracted mean CBF values extracted using the Yeo’s 7-network cortical atlas ROIs to examine network-level perfusion in the active and sham groups.

In addition, group-level CBF comparison of the whole brain was conducted using permutation-based inference (*randomise* in FSL) with threshold-free cluster enhancement (TFCE)^41^.

### 2.5. Statistical Analysis

Analyses were conducted in R v4.2.2. For each outcome, change scores (Δ = week 7 − baseline) were tested for normality using the Shapiro–Wilk test (α = 0.05). Between-group differences in change scores constituted the primary analyses; within-group effects are reported descriptively in the Supplementary Material. Normally distributed data were compared with independent t-tests (Welch’s correction if variances were unequal) and effect sizes expressed as Cohen’s d. Non-normal data were analyzed using Mann–Whitney U tests with rank-biserial correlation (r = Z/√N) as effect size. This framework was applied consistently across cognitive, biomarker, and imaging outcomes. Given the pilot nature and small sample size, no correction for multiple comparisons was applied. A priori hypotheses targeted mitochondrial and networklevel endpoints (lactate, pyruvate, lactate/pyruvate ratio, DMN FC); other outcomes were exploratory. Missing data were handled by pairwise deletion.

## 3. Results

### 3.1. Demographics

Participants were evenly split into active (n = 10) and sham (n = 10) groups. The mean age, sex distribution, and years of education did not differ significantly between the groups. Baseline MoCA scores were identical, confirming successful randomization.

### 3.2. Adherence and Safety

Adherence was high (active: 34.9 ± 2.0 sessions, 96.9%; sham: 33.9 ± 2.9 sessions, 94.2%), with 10/10 active and 9/10 sham participants completing ≥30 of 36 sessions. One sham participant discontinued after five weeks, believing the study had ended. No serious adverse events occurred; mild, self-limited events were reported by 10 of 20 participants (4/10 active; 6/10 sham), including transient scalp/head discomfort, headache or migraine, nasal/forehead dryness, low mood, or fatigue; all resolved spontaneously (Supplementary Table S1).

### 3.3. Cognitive Function

tPBM improved memory across multiple measures. CVLT-II long-delay recognition increased in the active group (+1.38 ± 2.39) but declined in sham (–2.00 ± 3.55), yielding a between-group difference of +3.38 (95% CI 0.44–6.32; t(16)=2.40, p=0.029, d=1.09). Long-delay cued recall similarly rose in active (+1.12 ± 1.95) and decreased in sham (–1.00 ± 2.30), difference +2.13 (95% CI –0.02–4.28; t(16)=2.11, p=0.051, d=0.98). Global cognition (MMSE) improved by +1.00 ± 1.25 in active versus –0.50 ± 1.58 in sham (+1.50; 95% CI 0.16–2.84; t(18)=2.36, p=0.035, d=1.05). Executive function (TMT-B) showed a trend favoring active treatment (– 32.26 ± 38.04 vs. –4.11 ± 23.85 s; p=0.066, d=–0.89). No significant effects were observed for processing speed (TMT-A), sleep (PSQI), or neuropsychiatric symptoms (MBI-C) (Figure 3).

**Figure 3.**
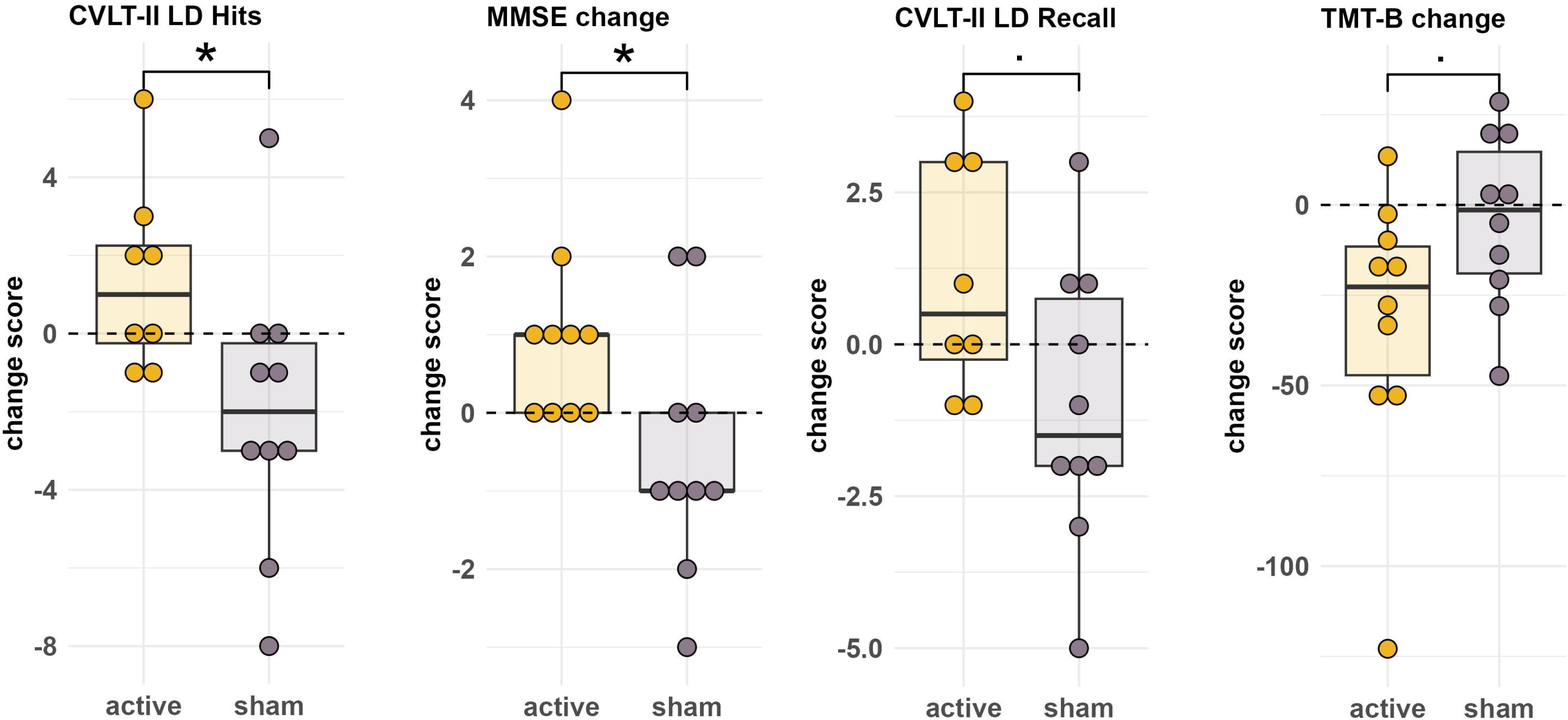
Changes in Cognitive Outcomes in the active group vs. sham group.

### 3.4. Blood-Based Biomarkers

#### 3.4.1. Serum Metabolomics

In the active group, serum pyruvate increased (+16.50 ± 28.44 µM) versus a decrease in sham (– 19.43 ± 23.09 µM), yielding a mean difference of +35.93 µM (95% CI 10.4–61.5; t(17)=3.04, p=0.0075, d=1.38). The lactate-to-pyruvate ratio decreased in active (–5.98 ± 12.58) but increased in sham (+8.67 ± 7.95), difference –14.65 (95% CI –25.0 to –4.3; t(17)=–3.06, p=0.0077, d=–1.37). Sarcosine rose less in active (+0.05 ± 1.03 µM) than in sham (+1.04 ± 0.89 µM), difference –0.99 (95% CI –1.91 to –0.07; t(17)=–2.24, p=0.038, d=–1.02). Urea decreased in active (–341.28 ± 585.56 µM) but increased in sham (+360.19 ± 771.41 µM), difference – 701.5 (95% CI –1376.6 to –26.4; t(17)=–2.21, p=0.042, d=–1.03). L-carnitine increased in active (+0.96 ± 10.18 µM) and declined in sham (–9.18 ± 10.33 µM), difference +10.14 (95% CI 0.2– 20.1; t(17)=2.15, p=0.046, d=0.99). L-alanine (+138.70 ± 195.82 µM vs –8.94 ± 122.29 µM; p=0.043, Z=2.04) and lactate (+417.70 ± 892.06 µM vs –183.56 ± 710.15 µM; p=0.035, Z=2.12) also rose significantly in the active group (Figure 4a).

**Figure 4.**
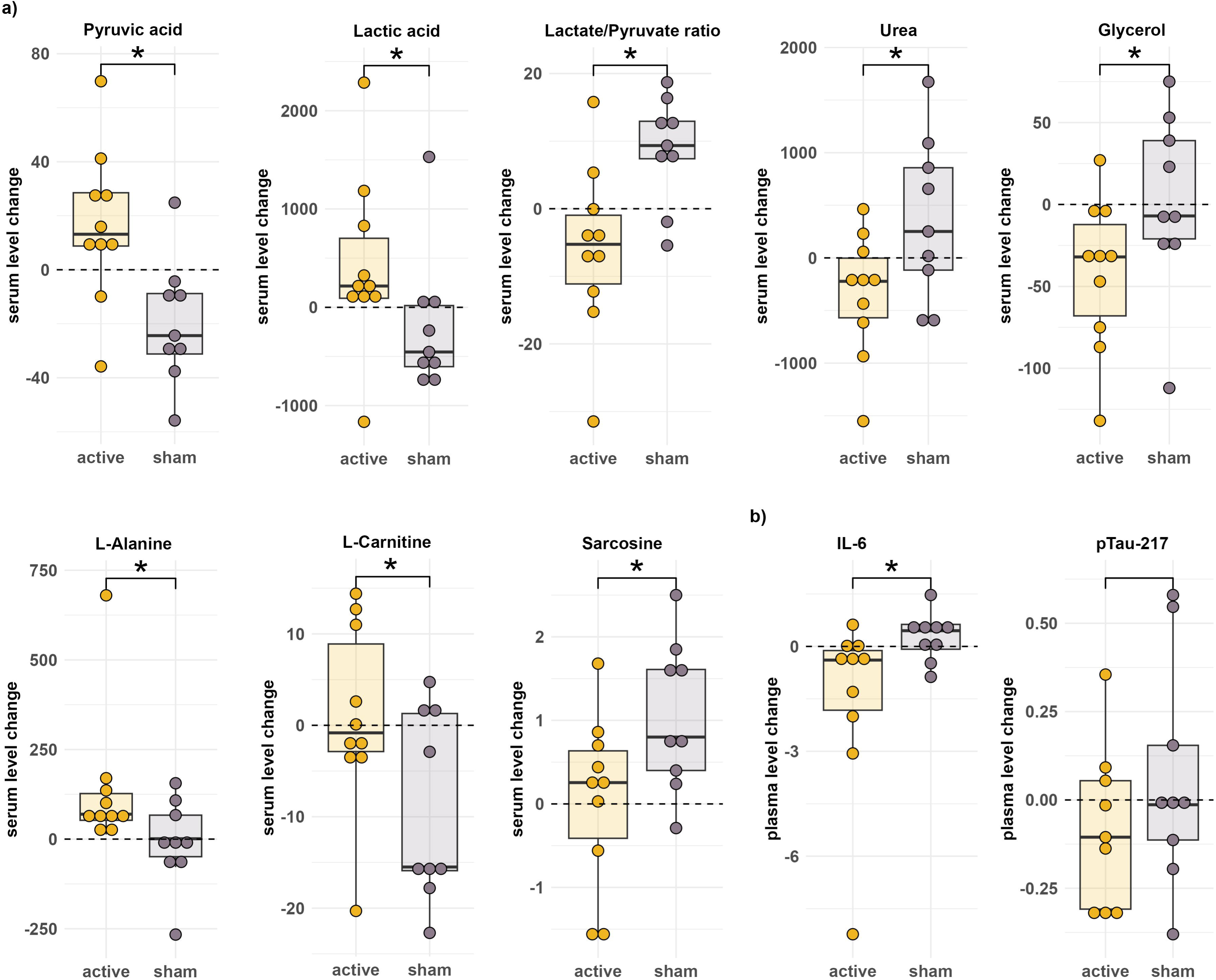
a) Changes in serum mitochondrial metabolites and related biomarkers in the active vs. sham group. b) Changes in plasma inflammatory marker (IL-6) in the active versus sham group.

#### 3.4.2. Plasma Biomarkers

Plasma IL-6 declined with active tPBM (–1.50 ± 2.61 pg/mL) but increased in sham (+0.27 ± 0.69 pg/mL), confirmed by a Wilcoxon test (W=17.5, Z=–2.24, p=0.027, r=–0.52; Hodges– Lehmann median difference –1.67 pg/mL, 95% CI –3.05 to –0.25) (Figure 4b). Plasma p-Tau217 levels did not differ significantly between the active and sham groups.

### 3.5. Neuroimaging

#### 3.5.1. rs-fMRI

Active tPBM increased DMN FC (+0.077 ± 0.126) compared with a decrease in sham (–0.095 ± 0.148), yielding a between-group difference of +0.172 (95% CI 0.041–0.303; t(17)=2.73, p=0.014, d=1.25). Caudate–DMN FC also increased in active (+0.057 ± 0.161) but declined in sham (–0.135 ± 0.166), difference +0.192 (95% CI 0.033–0.351; t(17)=2.55, p=0.021, d=1.17). Limbic network FC decreased in active (–0.149 ± 0.249) and rose in sham (+0.209 ± 0.079), difference –0.358 (95% CI –0.710 to –0.006; t(17)=–2.15, p=0.048, d=–0.99). No other networklevel effects were significant (Figure 5a).

**Figure 5.**
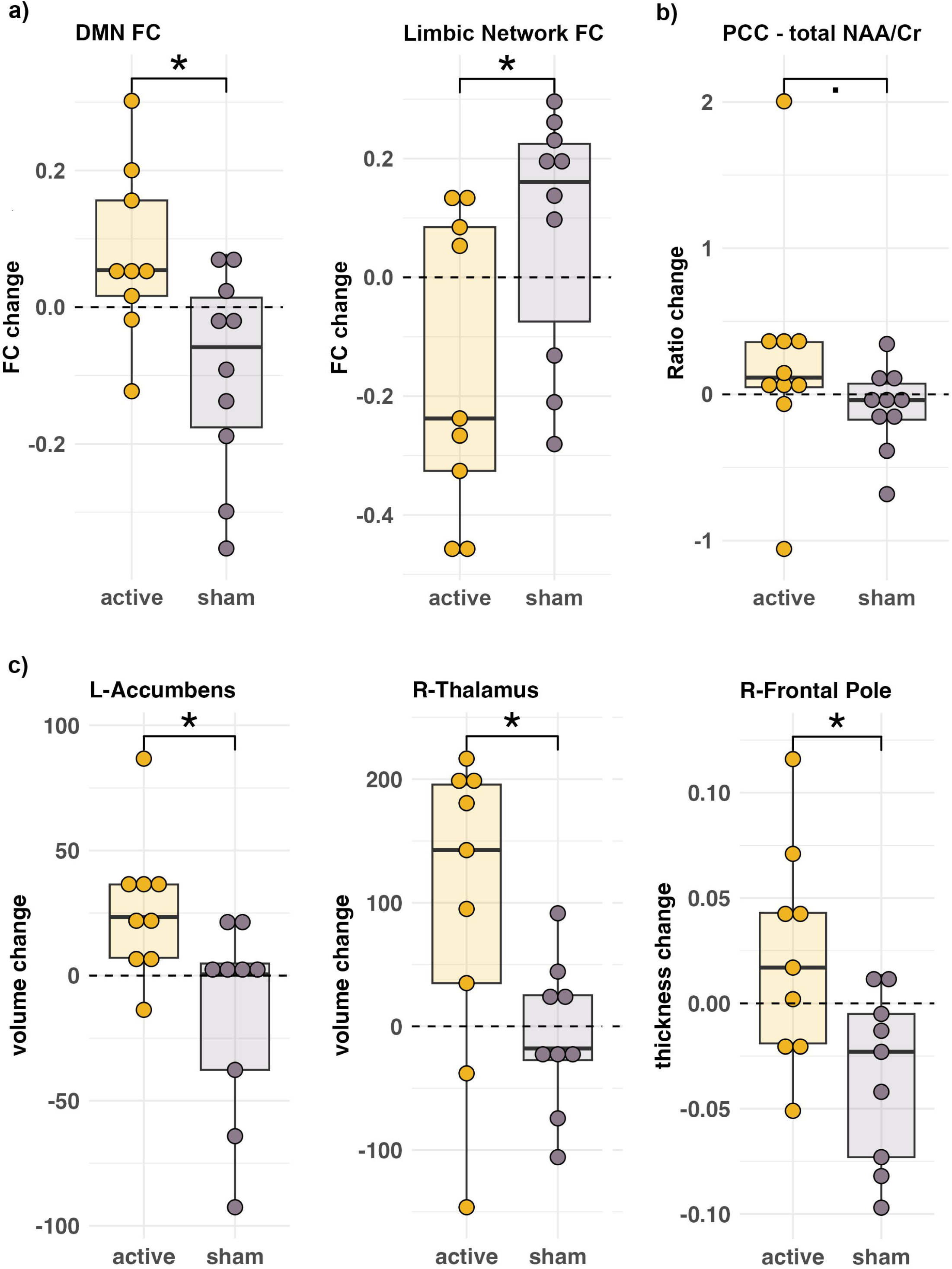
a) Functional connectivity changes with resting-state fMRI in the active versus sham group. b) Neurometabolite ratio changes in response to tPBM in the active versus sham group. c) Structural brain changes in the active versus sham group.

#### 3.5.2. ¹H-MRS

In the posterior cingulate cortex, the total N-acetylaspartate to creatine ratio (tNAA/Cr) trended higher in active (+0.230 ± 0.748) than sham (–0.091 ± 0.285), median difference +0.321 (95% CI –0.031–0.673; W=74, p=0.075, r=0.18). No other metabolites differed significantly (Figure 5b).

#### 3.5.3. Structural MRI - T1-Weighted Imaging

The left nucleus accumbens volume increased in active (+26.6 ± 28.3 mm³) but decreased in sham (–15.7 ± 40.1 mm³), difference +42.3 mm³ (95% CI 8.1–76.5; t(16)=–2.59, p=0.021, d=1.22). The right thalamus showed a similar pattern (+98.1 ± 125.2 mm³ vs –6.8 ± 60.5 mm³; 95% CI 3.2–206.6; t(16)=2.26, p=0.043, d=1.06).

Active tPBM increased cortical thickness in three right-hemisphere regions: frontal pole (+0.0568 mm, 95% CI 0.009–0.105; t(16)=2.58, p=0.021, d=1.22), entorhinal cortex (+0.1031 mm, 95% CI 0.013–0.193; t(16)=2.47, p=0.028, d=1.17), and pericalcarine gyrus (+0.0416 mm, 95% CI 0.005–0.078; t(16)=2.41, p=0.029, d=1.14), compared with decreases in the sham group (Figure 5c).

#### 3.5.4. Arterial Spin Labeling

No significant between-group differences in cerebral blood flow were detected.

## 4. Discussion

To our knowledge, this is the first sham-controlled randomized trial of transcranial photobiomodulation (tPBM) in Mild Cognitive Impairment (MCI) using a multimodal framework integrating cognition, blood-based metabolites, and neuroimaging biomarkers. Compared with sham, tPBM improved global cognition (MMSE) and episodic memory (CVLTII delayed recognition) and increased pyruvate and lactate with a reduced lactate-to-pyruvate ratio (L/P). Default mode network (DMN) functional connectivity (FC) also increased, indicating enhanced network-level brain function.

### 4.1. Hypothesis-Driven Outcomes

#### 4.1.1. Cognitive Function

The primary outcome was improvement in cognitive function. Global cognition improved significantly on the MMSE in the active group, in line with other studies^42^. The mean gain exceeded the 1-to 3-point threshold which is considered clinically meaningful^43^. Episodic memory also improved on the CVLT-II long-delay recognition task, showing more hits and fewer false positives. This is an especially meaningful finding given that recognition memory deficits emerge early in the ADRD trajectory (e.g., MCI)^44^.

#### 4.1.2. Serum Mitochondrial Biomarkers

Blood-based metabolic markers indicated a bioenergetic shift in the active tPBM group. Serum pyruvate rose more than lactate, lowering the L/P ratio and signaling greater oxidative phosphorylation through the tricarboxylic acid (TCA) cycle and increased mitochondrial engagement^45^. As MCI is often marked by elevated lactate^15^, normalization of the L/P ratio suggests partial restoration of cerebral energy metabolism. Because both metabolites cross the blood–brain barrier via monocarboxylate transporters^15^, serum levels provide accessible proxies of brain metabolism.

#### 4.1.3. Multimodal Neuroimaging Biomarkers

Proton magnetic resonance spectroscopy (¹H-MRS) showed a trend toward higher Nacetylaspartate/creatine (NAA/Cr) ratio in the posterior cingulate cortex (PCC), a key default mode network hub and early site of ADRD degeneration^16^. Although not significant, this trend may indicate improved neuronal viability or mitochondrial support^46^.

Resting-state fMRI showed increased DMN FC in the active tPBM group, suggesting enhanced communication between regions supporting memory, attention, and self-referential processing. The DMN was selected *a priori* for its central role in memory, early Alzheimer’s pathology^47^, and overlap with tPBM stimulation sites. This aligns with prior reports of tPBM-induced DMN FC increases in MCI^14^.

The absence of a significant change in cerebral blood flow (CBF) using pseudo-continuous arterial spin labeling (pCASL) may reflect its limitation in detecting transient or activity-dependent effects^48^. Prior PBM studies have shown short-lived CBF increases resolving within minutes to hours and measured by SPECT or other hemodynamic methods^49,50^. Alternatively, improved mitochondrial efficiency and oxygen utilization may reduce compensatory hyperperfusion, maintaining stable CBF despite enhanced oxidative metabolism^9,48^.

### 4.2. Exploratory Outcomes

#### 4.2.1. Extended Serum Metabolomics

Exploratory analyses revealed systemic metabolic effects of tPBM. Elevated L-alanine suggests increased pyruvate transamination^45^, and higher L-carnitine indicates enhanced mitochondrial fatty acid import and β-oxidation^51^. Reduced urea and sarcosine imply decreased amino acid catabolism and altered NMDA signaling^52^, while lower glycerol reflects reduced lipolysis^53^. Along with higher pyruvate and lactate and a lower lactate-to-pyruvate ratio, these findings indicate improved aerobic ATP production and metabolic efficiency.

#### 4.2.2. Plasma Inflammatory Biomarkers

Active tPBM lowered plasma IL-6, a cytokine linked to systemic inflammation and AD progression^31^. This is consistent with in vitro evidence of PBM-mediated IL-6/IL-8 suppression and clinical trials showing reduced IL-6, IL-8, and TNF-α in COVID-19 patients^54^, supporting the hypothesis that tPBM attenuates inflammation via mitochondrial mechanisms^9^.

#### 4.2.3. Exploratory Neuroimaging

Exploratory rs-fMRI revealed increased caudate–DMN FC, suggesting enhanced network integration for memory and cognitive control^55^, and decreased limbic connectivity, potentially reflecting reduced emotional reactivity, often heightened in early AD^16^.

Structural MRI showed volume increases in the left nucleus accumbens and right thalamus, regions associated with motivation, attention, and executive control that typically exhibit early AD atrophy^16^. Modest cortical thickening in the right frontal pole and entorhinal cortex—areas central to working and episodic memory^16^—further supports functional improvement. Similar short-term gray matter and cortical thickness gains have been observed after 2–6 weeks of highfrequency rTMS, reflecting rapid, activity-dependent neuroplasticity^56,57^. Given the comparable timeframe, the present subcortical changes are more plausibly physiologic or metabolic, reflecting modulation of perfusion and mitochondrial activity rather than large-scale structural remodeling. Thus, these volumetric effects are interpreted as functional or metabolic adaptations rather than morphologic growth.

### 4.3. Unique Aspects of the Intervention

A distinguishing feature of this protocol was intranasal LED stimulation targeting the olfactory bulb, an early site of ADRD pathology^58^. This route may engage subcortical and limbic circuits via olfactory projections^58^, enhancing downstream neuromodulation. Monte Carlo modeling and optical data indicate that only 1–3% of 810-nm light penetrates the scalp and skull (∼0.75–3 mW/cm²)^20,59^, whereas the intranasal emitter bypasses this barrier to illuminate the olfactory epithelium and bulb. These structures connect with limbic and prefrontal regions of the default mode network (DMN), which shows early metabolic decline in MCI^47^. The observed cortical and subcortical effects may therefore reflect indirect modulation of this olfactory–limbic–prefrontal circuit through metabolic and network-level mechanisms.

### 4.4. Mechanistic Synthesis of Findings

This study provides preliminary evidence that tPBM modulates cognition and physiological processes relevant to early neurodegeneration. Increases in serum pyruvate and lactate, a lower lactate-to-pyruvate ratio, higher L-carnitine and L-alanine, and reduced urea, glycerol, and sarcosine indicate enhanced oxidative metabolism with diminished reliance on amino acid and lipid catabolism. Concurrent IL-6 reduction suggests anti-inflammatory effects, while increased DMN FC reflects greater stability in memory-related circuits. The intranasal LED component may augment these effects by engaging olfactory–limbic pathways.

Although neurovascular coupling generally links oxidative metabolism and cerebral blood flow (CBF), the relationship is nonlinear and modulated by mitochondrial efficiency and oxygen buffering^60^. PBM likely enhances oxidative phosphorylation^9^, allowing increased metabolism without measurable CBF change.

Together, these findings support a model in which tPBM promotes cognitive benefits through mitochondrial and immunometabolic modulation that influences large-scale brain networks.

### 4.5. Strength and Limitations

Strengths include the randomized, sham-controlled design; integration of cognition, blood-based, and multimodal neuroimaging biomarkers provided mechanistic insight; and visually indistinguishable devices and high adherence with indistinguishable sham devices, supporting feasibility and internal validity. Limitations include the small sample size (n=10/group), which reduced statistical power and precluded correction for multiple comparisons. Because a single unblinded coordinator administered cognitive assessments, expectancy bias cannot be excluded; future studies will employ full double-blinding with independent assessors and concealed allocation. PET or CSF biomarkers were not collected to confirm underlying ADRD pathology. Although participants were instructed to use the device at bedtime to standardize session timing, variability in the exact timing of use and participants’ pre-treatment state may have contributed to outcome variability and should be more tightly controlled in future trials. In addition, because near-infrared light penetration through the scalp and skull is limited (∼1–3%), restricting direct illumination of deep midline regions, the observed network-level effects likely reflect indirect modulation of default-mode and limbic pathways through cortical and olfactory projections rather than direct irradiation. Finally, durability of effects beyond the treatment window was not assessed. These findings should therefore be considered preliminary and hypothesis-generating.

## 5. Conclusion

This feasibility trial shows that home-based tPBM is safe, well-tolerated, and demonstrates preliminary efficacy across cognitive and biomarker outcomes in MCI. The findings support a mitochondrial-targeted, noninvasive approach capable of modulating brain structure, connectivity, and systemic metabolism. As anti-amyloid agents like lecanemab enter clinical use, tPBM may provide complementary benefits by addressing non-amyloid mechanisms. Overall, tPBM emerges as a promising candidate for early ADRD intervention, warranting larger doubleblind, multicenter trials with longitudinal cognitive endpoints.

## Acknowledgments

We gratefully acknowledge TDRA-MITO2i for awarding a fellowship to the first author, and Temerty-Tanz-TDRA for their seed funding, which enabled the inception of this work. We also thank Hilary and Galen Weston Foundation for generously funding the study. We are grateful to Janine Liburd and Genane Loheswaran for their technical support and assistance in responding to device-related questions throughout the trial. Finally, we also acknowledge the patients who participated in the study and their families without which this work could not have been completed.

## Consent Statement

All study participants provided written informed consent.

## Data Availability

The data that support the findings of this study are available from the corresponding author upon reasonable request.

## Code Availability

Code for statistical analyses and figure generation, including all data processing scripts, is available at: https://github.com/NedaRR/PBMCI_notebook.git. Code for the MRI processing supporting this study will be made available upon reasonable request.

## Contributions

NRR conceived and designed the study, obtained ethics approval, conducted cognitive assessments, processed MRI data, performed all statistical analyses for cognitive, blood-biomarker and MRI data, created the figures and wrote the manuscript.

NWC optimized MRI sequences, provided processing pipelines for resting-state fMRI and arterial spin labeling, and reviewed the manuscript. MJ centrifuged and aliquoted blood samples, performed Ella assays, and reviewed the manuscript. RS supported Quanterix plasma biomarker assays, and reviewed the manuscript. RZ provided photobiomodulation expertise, assisted with methodological design, and reviewed the manuscript. OR supported the Ella assays and reviewed the manuscript. ACA advised on mitochondrial assays, contributed to data interpretation, and reviewed the manuscript. TAS supported MRI acquisition and data-processing workflow and reviewed the manuscript. TKR, SJG, DGM, LF, and MN provided scientific guidance and reviewed the manuscript. LL supplied photobiomodulation devices and reviewed the final version of the manuscript to verify the technical accuracy of the device description. CEF codesigned the study, recruited participants, supervised trial conduct, and reviewed the manuscript (corresponding author).

## Funding

This work was supported by the Toronto Dementia Research Alliance–Mitochondrial Innovation Initiative (TDRA–MITO2i) Fellowship, awarded to the first author (NRR) for this project; the Hilary and Galen Weston Foundation, which funded the clinical trial; and the TDRA Seed Fund, which supported the biomarker analyses.

## Competing interests

NRR has received consulting fees from Vielight Inc. (2021 – 2023). RZ has received consulting fees from Vielight Inc. (2018 – present). RS holds the Elizabeth S. Barford Early Career Professor in Multiple Sclerosis in the Department of Medicine at the University of Toronto. He has received a Discovery Grant from MS Canada and additional funding from Brain Canada to study the effects of Epstein-Barr Virus on Multiple Sclerosis. He has received consulting fees from Novartis and EMD Serono and payments or honoraria for lectures, presentations, and educational events from Biogen-Idec, Sanofi-Genzyme, EMD Serono, Roche and Eli Lilly. RS has participated on advisory boards for Novartis and EMD Serono. He has also received support for attending scientific meetings from EMD Serono. LL is the Chief Executive Officer of Vielight Inc. and provided the study devices. LL’s involvement was limited to verifying the technical accuracy of the device description. All raw data and code resided on institutional servers; analyses were performed by academic investigators; the device manufacturer had no access to interim results. CEF has received research grants from Hoffmann-La Roche (2018 – 2022), Vielight Inc. (2019 – 2023) and Novo Nordisk (2021 – 2024). The remaining authors declare no competing interests.

## Notes

### Clinical Trial

NCT05563298

### Clinical Protocols

https://www.clinicaltrials.gov/study/NCT05563298

### Author Declarations

REB# 22-128 - A pilot study evaluating the feasibility, safety, and efficacy of the Vielight Neuro RX Gamma for the treatment of amnestic MCI REB APPROVAL: Original Approval Date February 09, 2023 Annual/Interval Review Date February 09, 2024 Thank you for your application submitted on May 30, 2022. At the Unity Health Toronto Research Ethics Board (REB) meeting held on June 29, 2022, the above referenced study was discussed and subsequently the views derived from this discussion have been documented and resolved. Please note that no member of the REB associated with this study was present or involved in its deliberation, review or approval. The REB approves the study as it is found to comply with relevant research ethics guidelines, as well as the Ontario Personal Health Information Protection Act (PHIPA), 2004. The REB hereby issues approval for the above named study for a period of 12 months from the date of this letter. Continuation beyond that date will require further review of REB approval.

### Summary of Updates

The adherence and adverse-event data presented in the revised manuscript reflects source-verified entries from participant diaries and coordinator call notes. This review resulted in minor numerical adjustments but did not alter any study outcomes or conclusions.

